# The Interplay between Rapid Mental Health Screening and Oral Health Behaviors – A Population-Based study with a Focus on Toothbrushing Frequency and Dental Attendance

**DOI:** 10.1101/2025.10.24.25338743

**Authors:** Shabnam Varmazyari, Keyvan Karimi, Yosra Azizpour, Samaneh Akbarpour, Mojgan Alaeddini

## Abstract

**Objectives:** Little is known about how major mental distresses such as depression, anxiety, and suicide impact key oral health behaviors, particularly in low- and middle-income countries (LMICs) like Iran. This study assesses whether depressive and anxious moods and suicidal ideation, rapidly screened indicators of these distresses, relate to toothbrushing frequency and dental attendance among a sample of Iranian adults.

**Methods:** This cross-sectional study used data from the Integrated and Repeated Public Health Surveillance project (May–September 2023) in southern Tehran Province, Iran. Stratified random sampling was used to survey adults aged ≥18 via telephone. A validated questionnaire assessed depressive mood, anxious mood, suicidal ideation, toothbrushing frequency, and 6-month dental attendance. A “composite mental distress” variable was also created by combining the three mental distresses. Binary logistic regression analyzed associations, adjusting for covariates.

**Results:** Among the 1,282 participants (mean age 40.1, SD = 13.3, 60.6% female), 23.5% toothbrushed twice a day and above, and 33.2% attended dental visits in the past 6 months. Moderate and high depressive mood levels, higher depressive mood scores, and higher anxious mood scores were all related to toothbrushing less than twice daily (AOR = 0.65, p = 0.004, AOR= 0.54, p = 0.037, AOR = 0.85, p = 0.006, and AOR = 0.87, p = 0.021, respectively). Moderate composite mental distress levels and higher composite mental distress scores were also related to toothbrushing less than twice daily (AOR = 0.52, p < 0.001, and AOR: 0.94, p **=** 0.019, respectively). None of the mental distress indicators were associated with 6-month dental attendance.

**Conclusions:** To address findings, oral healthcare providers should be trained in rapid mental health screening, provide toothbrushing support for those screened, and refer them to mental healthcare when necessary. Similarly, mental healthcare providers should be trained to recognize mental distress indicators of poor toothbrushing, promote toothbrushing among therapeutic goals, and refer patients to oral healthcare when necessary

## Introduction

Oral health, the ability to taste, chew, swallow, smile, speak, and express emotions without pain, discomfort, or disease in the craniofacial region [1], is an essential aspect of overall health and well-being, with strong associations to diabetes, cardiovascular and respiratory diseases, metabolic syndromes, and Alzheimer’s disease [2–4]. However, the reductionist biomedical model has long separated oral health from general health, leading to higher treatment costs, more severe health inequities, and fragmented care [4, 5]. In contrast, the biopsychosocial model and integrated care frameworks emphasize that oral health is inseparable from overall well-being, reflecting shared biological, behavioral, and social determinants [6, 7]. These perspectives remain insufficiently implemented in low- and middle-income countries (LMICs), where the repercussions of neglecting oral health integration are more pronounced [8, 9]. Iran is one such country with high oral disease burden, limited financial and infrastructural resources, and poor integration of oral health within the general healthcare system [10–13].

Mental healthcare is no exception to the systemic separation from oral health. Providers in these two domains often work independently, receive little cross-disciplinary training, and have limited time or resources for collaboration [14, 15]. This disconnect is concerning, as major mental distresses, such as depression, anxiety, and suicide, can profoundly affect oral health. Depression and anxiety, which together affect over 690 million people worldwide and cost the global economy an estimated US$1 trillion annually [16, 17], as well as suicide, which claims more than 700,000 lives each year [18], heighten vulnerability to temporomandibular disorders, xerostomia, and oral ulcerations; increase the risk of periodontal disease, dental caries, and tooth loss; and impair chewing, speech, and overall oral function [19–28].

Behavioral pathways may partly explain the associations between these mental distresses and oral health. Twice-daily toothbrushing and regular dental attendance, two key oral health behaviors, play a crucial role in preventing plaque accumulation, dental caries, periodontal disease, and tooth loss [29–31]. Several studies have reported associations between the mental distresses of depression, anxiety, and suicide infrequent toothbrushing, and irregular dental attendance [23, 32–36]. However, the evidence remains inconsistent, as some studies have found no association or even opposite trends between these mental distresses, toothbrushing frequency, and dental attendance [22, 37–39]. Moreover, much of the existing research has focused narrowly on dental anxiety rather than broader anxiety constructs [20], treated suicide as part of composite indices instead of an independent construct [36, 40, 41], relied on outdated data [33, 42], or been conducted within specific contexts and populations, such as during the COVID-19 pandemic or among airline pilots [25, 32]. Notably, no studies to date have specifically examined the association between suicidal ideation, attempts, or behavior and dental attendance. Furthermore, Iranian adults remain largely underrepresented in this area of research.

To help bridge the divide between mental and oral healthcare and address these research gaps, timely detection of these mental distresses in oral healthcare settings is essential [14, 15, 43]. Instruments such as the Patient Health Questionnaire-9 (PHQ-9), PHQ-4, the Generalized Anxiety Disorder-7 (GAD-7), the World Health Organization Well-Being Index, and the General Health Questionnaire-28 (GHQ-28) offer efficient means of identifying mental distress in busy oral healthcare environments [44–47]. Because these tools are brief, valid, and reliable, they can effectively capture the core symptoms of each distress and flag individuals in need of referral and further assessment [44–47]. However, because rapid screening tools are primarily designed for population-level surveillance and triage, they are better suited for assessing current emotional states and thoughts rather than providing formal diagnoses. Accordingly, this study uses them to evaluate depressive and anxious moods rather than full clinical diagnoses of depression or anxiety disorders. Additionally, as suicidal ideation is an early warning sign of suicidal behavior [18], rapid screening is employed to assess this construct rather than actual suicidal behavior. Therefore, the present study examines whether the rapidly assessed mental distress indicators of depressive mood, anxious mood, and suicidal ideation are associated with toothbrushing frequency and dental attendance among a sample of Iranian adults.

## Methods

### Ethical considerations

This study was carried out in compliance with the Helsinki declaration. Verbal informed consent was obtained prior to study conduction, after informing participants of the study’s purpose, the voluntary nature of their involvement, anonymity preservation, data confidentiality, and the estimated duration of 15–20 minutes. Ethical approval was also obtained prior to commencing the study by the Research Ethics Committee of school of medicine at Tehran University of Medical Sciences (TUMS) (ethics code: IR.TUMS.MEDICINE.REC.1400.599).

### Study Design and Setting

This study draws on data from the Integrated and Repeated Public Health Surveillance (IRPHS) project, a population-based telephone survey employing a cross-sectional design. The methodological process of the IRPHS is detailed in its published protocol [48]. Here, only a brief summary of the most important points is provided.

### Target population and sampling strategy

The IRPHS targeted adults aged 18 years and older residing in areas supervised by TUMS, which include southern Tehran city, Eslamshahr, and Shahr-e Rey. These areas represent one of three administrative regions within Tehran Province, each overseen by a separate medical university.

Stratified random sampling with probability proportional to size was used to ensure representativeness across the three TUMS-supervised strata. Sample size calculations were conducted separately for each major health indicator using the population proportion formula and 2021 Iranian STEPS data [49]. The largest required sample was 1,199 individuals, inflated to 1,332 to account for a 10% attrition rate. Within each stratum, household phone numbers were randomly selected, and one eligible adult per household was interviewed. Eligibility required the respondent to be a family member who had resided in the household for at least six months and was expected to remain for the following month.

### Data Collection Instrument

The IRPHS monitored key health indicators selected through expert consultation and scientific evidence. These included demographic characteristics, healthcare utilization, chronic disease prevalence, lifestyle behaviors, and mental and oral health status. A structured questionnaire was developed to capture these domains. Content validity was assessed using the Content Validity Ratio (CVR ≥ 0.59) and Content Validity Index (CVI ≥ 0.79); items falling below these thresholds were revised or excluded. Reliability was confirmed with an agreement coefficient ≥ 0.7.

### Interviewer Training and Quality Assurance

Interviewers with prior survey experience underwent targeted training. To ensure data integrity, pilot interviews were conducted, calls were recorded and randomly reviewed, and 10% of completed questionnaires were independently rechecked. A subset of responses was validated through face-to-face interviews and clinical assessments. Data collection occurred between May 22 and September 24, 2023. Upon completion of interviews, participants identified as at-risk were referred to local health centers for free physician consultations and informed about additional support services available within the TUMS-supervised region.

### Study measures

#### Outcome Variables

Toothbrushing frequency: Participants were asked a single question about how often they brush their teeth, with the response options: “Three times a day,” “Twice a day,” “Once a day,” “Once a week,” “Two to three times a week,” “Never,” and “I wear complete dentures.” Participants who wore complete dentures were excluded. For descriptive analysis, responses were grouped into three categories: “At least twice a day,” “Once a day,” and “Less than once a day.” For logistic regression, the responses were further dichotomized into a binary variable, following similar categorizations in the literature [23, 41], and aligning with international guidelines for minimum daily brushing [50]. The final categories were “Twice a day and above” and “Less than twice a day”, which combined all other responses.

Six-month dental visit attendance: Participants were asked a single question about whether they had visited a dentist in the past six months, with responses of “No” or “Yes.” The 6-month timeframe was chosen to capture dental visits for several reasons. First, previous studies have successfully used this period, as shown in a scoping review of adult dental care utilization [51]. Second, shorter recall periods, such as 6 months instead of 12, improve the accuracy of self-reported health service use [52, 53]. Finally, while salient events like hospitalizations can be remembered over longer periods, frequent routine visits are more easily forgotten, making a 6-month recall reliable [54].

#### Independent Variables

Depressive mood: Depressive mood was assessed using the two PHQ-2 questions, validated in Persian [55], asking about lack of interest in usual activities and feelings of depression or hopelessness over the past two weeks. Responses were scored from 0 (”Not at all”) to 3 (”Nearly every day”) and summed to create a depressive mood score (0–6), with higher scores indicating greater severity. Scores were divided into tertiles to form low (0–1), moderate (2–3), and high (4–6) depressive mood levels.

Anxious mood: Anxious mood was assessed using two questions adapted from the GAD-2 and validated in Persian [56, 57], asking how often participants felt nervous or anxious and how well they could control their worries over the past two weeks. Responses ranged from 0 (”Not at all”) to 3 (”Nearly every day”) and were summed to create an anxious mood score (0–6), with higher scores indicating greater anxiety. Scores were also divided into tertiles to form low (0–1), moderate (2–3), and high (4–6) anxious mood levels.”

Suicidal ideation: Suicidal ideation was assessed using two questions adapted from the GHQ-28 and validated in the IRPHS [49, 58, 59], asking about feelings of hopelessness and thoughts of ending one’s life over the past month. Responses ranged from 0 (”None or minimal”) to 3 (”Severe”) and were summed to create a suicidal ideation score (0–6), with higher scores indicating greater severity. A 1-month timeframe was used to align with the GHQ-28, match the 2-week measures for depressive and anxious mood, and improve sensitivity given the low prevalence of suicidal ideation in the sample. For analysis, scores were dichotomized into low (0–1) and high (2–6), resulting in the variables “low suicidal ideation level” and “high suicidal ideation level”.

Composite mental distress: To capture the combined burden of depressive mood, anxious mood, and suicidal ideation, this study drew on similar constructs in the literature, such as “psychological distress” or “poor mental health” [23, 40, 41], to create a composite mental distress variable. Scores for depressive mood, anxious mood, and suicidal ideation were summed to generate this variable, which ranged from 0 to 18, with higher values indicating greater mental distress. The composite scores were also divided into tertiles (low, moderate, high) based on the sample distribution.

### Covariates

Covariates for this study were selected from variables consistently adjusted for in the literature on mental health and oral health behaviors [22, 36, 60, 61]. The covariates and their response options were: age (years); sex (male, female); education level (no education [0 years], primary [1–6], secondary [7–12], higher [>12]); residential area (urban, rural); socioeconomic status (high, above average, average, below average, low); current tobacco smoking (yes, no); marital status (single, married, divorced/widowed); chronic disease status (none, one, or two or more of hypertension, diabetes, cardiovascular disease, or dyslipidemia); and health service use in the past year (yes, no).

### Statistical analysis

Descriptive statistics included frequency and percentage for categorical variables. Differences in variable levels across toothbrushing frequency and 6-month dental attendance subgroups were assessed using chi-square or Fisher’s exact tests, as appropriate. Analysis of variance (ANOVA) was used to compare mean scores for depressive mood, anxious mood, and suicidal ideation, and mental distress across toothbrushing and 6-month dental attendance. Binary logistic regression examined the associations between mental distress and its components (as exposures) and toothbrushing frequency and 6-month dental attendance (as outcomes). Crude logistic regression models were first conducted for each mental distress variable and outcome, followed by adjusted models controlling for the covariates. Results were reported as odds ratios (ORs), adjusted odds ratios (AORs), and 95% confidence intervals (CIs). Data analysis was performed using Stata version 17, with a significance level set at p < 0.05. Cases with missing data on outcome variables (n=29) were excluded from analysis.

### AI declaration

In preparing the present manuscript, the authors used ChatGPT (OpenAI, San Francisco, CA, USA) for assistance with text editing, including improving readability and conciseness. All content was reviewed and verified by the authors for accuracy and integrity.

## Results

The study included 1,282 participants, as shown in Table 1. The overall population was predominantly female (60.6%), with a mean age of 40.1 years (SD = 13.3). Most participants (58.4%) were between 26 and 45 years old, and the largest educational subgroup (35.5%) had 7 to 12 years of schooling. The sample was predominantly urban (89.5%) and largely comprised of married individuals (78.9%). A significant portion (56.1%) reported poor SES.

**Table 1.** Toothbrushing Frequency and 6-month Dental Attendance Prevalence by Sociodemographics among Tehran Province Adults (n=1282).

| Variable | Variable category<br>N [%]<br>in the overall population | Tooth Brushing Frequency<br>N [%] in each subgroup |  |  |  |  | 6-month dental attendance<br>N [%] in each subgroup |  |  |
| --- | --- | --- | --- | --- | --- | --- | --- | --- | --- |
|  |  | Not At all<br>104 [8.1] | Sometimes (weekly)<br>106 [8.3] | Once a day<br>771 [60.1] | Twice a day<br>and above<br>301 [23.5] | p-value | Yes<br>425 [33.2] | No<br>856 [66.8] | p-value |
| Sex | Male<br>505 [39.4] | 61 [58.6] | 61 [57.5] | 326 [42.3] | 57 [18.9] | <0/001 | 127 [25.2] | 377 [74.8] | <0/001 |
|  | Female<br>777 [60.6] | 43 [41.3] | 45 [42.4] | 445 [57.7] | 244 [81.1] |  | 298 [38.3] | 479 [61.6] |  |
| Age (year) | 18-25<br>154 [12.0] | 6 [5.8] | 12 [11.3] | 94 [12.19] | 42 [13.9] |  | 56 [36.4] | 98 [63.6] |  |
|  | 26-45<br>749 [58.4] | 18 [17.3] | 65 [61.3] | 465 [60.3] | 201 [66.8] | <0/001 | 271 [36.2] | 477 [63.7] | <0/001 |
|  | 46-65 | 52 [50.0] | 22 [20.7] | 198 [25.7] | 50 [16.6] |  | 93 [28.9] | 229 [71.1] |  |
|  | 322 [25.12] |  |  |  |  |  |  |  |  |
|  | **>65**<br>57 [4.4] | 28 [26.9] | 7 [6.6] | 14 [1.8] | 8 [2.7] |  | 5 [8.77] | 52 [91.2] |  |
| **Residency** | **Urban**<br>1148 [89.5] | 88 [84.6] | 90 [84.9] | 688 [89.2] | 282 [93.7] | 0.013 | 388 [33.8] | 760 [66.2] | 0.17 |
|  | **Rural**<br>134 [10.4] | 16 [15.4] | 16 [15.1] | 83 [10.8] | 19 [6.3] |  | 37 [27.8] | 96 [72.2] |  |
| **Education** | **0 year**<br>241 [18.9] | 63 [60.6] | 42 [39.6] | 114 [14.8] | 22 [7.5] |  | 42 [17.4] | 199 [82.5] |  |
|  | **1-6 year**<br>228 [17.9] | 19 [18.3] | 21 [19.8] | 149 [19.3] | 39 [13.3] | <0/001 | 71 [31.0] | 158 [69] | <0.001 |
|  | **7-12 year**<br>453 [35.5] | 15 [14.4] | 28 [26.4] | 278 [36.1] | 132 [44.9] |  | 156 [34.4] | 297 [65.5] |  |
|  | **12< years**<br>353 [27.7] | 7 [6.7] | 15 [14.1] | 230 [29.8] | 101 [34.3] |  | 153 [43.3] | 200 [56.7] |  |
| **Marital status** | **Single** | 8 [7.7] | 13 [12.3] | 117 [15.2] | 56 [19.0] |  | 74 [38.1] | 120 [61.9] | 0.005 |
|  | 194 [15.2] |  |  |  |  |  |  |  |  |
|  | **Divorced/widow**<br>75 [5.9] | 17 [16.3] | 8 [7.5] | 34 [4.4] | 16 [5.4] | <0/001 | 13 [17.3] | 62 [82.7] |  |
|  | **Married**<br>1006 [78.9] | 79 [76.0] | 85 [80.2] | 620 [80.4] | 222 [75.5] |  | 335 [33.3] | 672 [66.7] |  |
| **SES** | **Poor**<br>712 [56.1] | 91 [87.5] | 56 [53.3] | 461 [60.0] | 104 [35.7] | <0/001 | 222 [31.2] | 490 [68.8] |  |
|  | **Moderate**<br>494 [39.0] | 11 [10.6] | 45 [42.9] | 277 [36.1] | 161 [55.3] |  | 170 [34.4] | 324 [65.6] | 0.03 |
|  | **Good**<br>62 [4.9] | 2 [1.9] | 4 [3.8] | 30 [3.91] | 26 [80.9] |  | 29 [46.8] | 33 [53.2] |  |
| **Health care visits<sup>#</sup>** | **No**<br>644 [50.2] | 73 [70.2] | 40 [37.7] | 383 [49.7] | 148 [49.2] | <0.001 | 209 [32.5] | 434 [67.5] | 0.607 |
|  | **Yes**<br>638 [49.8] | 31 [29.8] | 66 [62.3] | 388 [50.3] | 153 [50.8] |  | 216 [33.9] | 422 [66.1] |  |
| **Smoking status<sup>1</sup>** | **No** | 74 [71.1] | 81 [76.4] | 628 [81.6] | 260 [86.38] | 0.031 | 359 [34.42] | 684 [65.58] | 0.050 |
|  | 1043 [81.4] |  |  |  |  |  |  |  |  |
|  | **Yes**<br>238 [18.6] | 30 [28.9] | 25 [23.6] | 142 [18.4] | 41 [13.6] |  | 66 [27.8] | 171 [72.1] |  |
| **Chronic disease**<br>**status\*\*** | **None**<br>1044 [81.4] | 57 [54.8] | 83 [78.3] | 650 [84.3] | 254 [84.4] |  | 348 [33.4] | 695 [66.6] |  |
|  | **One**<br>162 [12.6] | 13 [12.3] | 88 [11.4] | 35 [11.6] | 13 [12.26] | <0.001 | 59 [36.4] | 103 [63.6] | 0.144 |
|  | **Two and above**<br>76 [5.9] | 21 [20.2] | 10 [9.4] | 33 [4.3] | 12 [3.99] |  | 18 [23.7] | 58 [76.3] |  |

The distribution of oral health behaviors varied considerably across demographic subgroups, as also shown in Table 1. Among participants who brushed their teeth twice a day and above, the vast majority were female (81.1%). A similar gender gap was observed in dental attendance within the past 6 months, with attendance highest among females (38.3%) and lowest among males (25.2%) (p < 0.001). Age differences were also striking. The largest share of those who brushed their teeth twice a day and above was found in the 26–45 age group (66.8%), while the smallest proportion came from those over 65 years (2.7%). For 6-month dental attendance, young adults aged 18–25 represented the largest group (36.4%), whereas older adults over 65 again reported the lowest rate (8.8%) (p < 0.001). Educational attainment also showed a gradient. Among participants who brushed their teeth twice a day and above, individuals with 7–12 years of schooling had the highest proportion (44.9%), compared with only 7.48% of those with no formal education. Similarly, for attending dental visits within 6 months, the largest proportion was seen among those with more than 12 years of schooling (43.3%), while the lowest was among individuals with no formal education (17.4%) (p < 0.001). SES was also strongly associated with oral health behaviors. The prevalence of toothbrushing twice a day and above was lowest among those in the poor SES group (35.7%) and highest among those in the good SES group (80.9%). A comparable pattern was observed for 6-month dental visits, where only 31.2% of participants from the poor SES group attended, compared to 46.8% in the good SES group.

As shown in Table 2, toothbrushing frequency was significantly associated with depressive (p=0.002) and anxious (p=0.010) mood scores, with lower scores for both mental distress indicators corresponding to higher toothbrushing frequency. Participants with low depressive mood scores brushed their teeth twice a day and above had a higher prevalence (53.9%) than those with high depression scores (7.2%, p=0.005). Similarly, low anxious mood levels were associated with a higher frequency of toothbrushing (58.1% for low anxious mood vs. 12.0% for high anxious mood, p=0.003). Categorical analysis of composite mental distress was also associated with more frequent toothbrushing (p<0.001), with high levels of mental distress related to lower toothbrushing frequency (50% for twice daily or more vs. 26.2% for not at all). Suicidal ideation was not associated with toothbrushing (p = 0.840) or dental attendance in the past 6 months (p = 0.458). Additionally, no associations were found between 6-month dental attendance and any of the mental distress indicators (all p > 0.05).

**Table 2.**
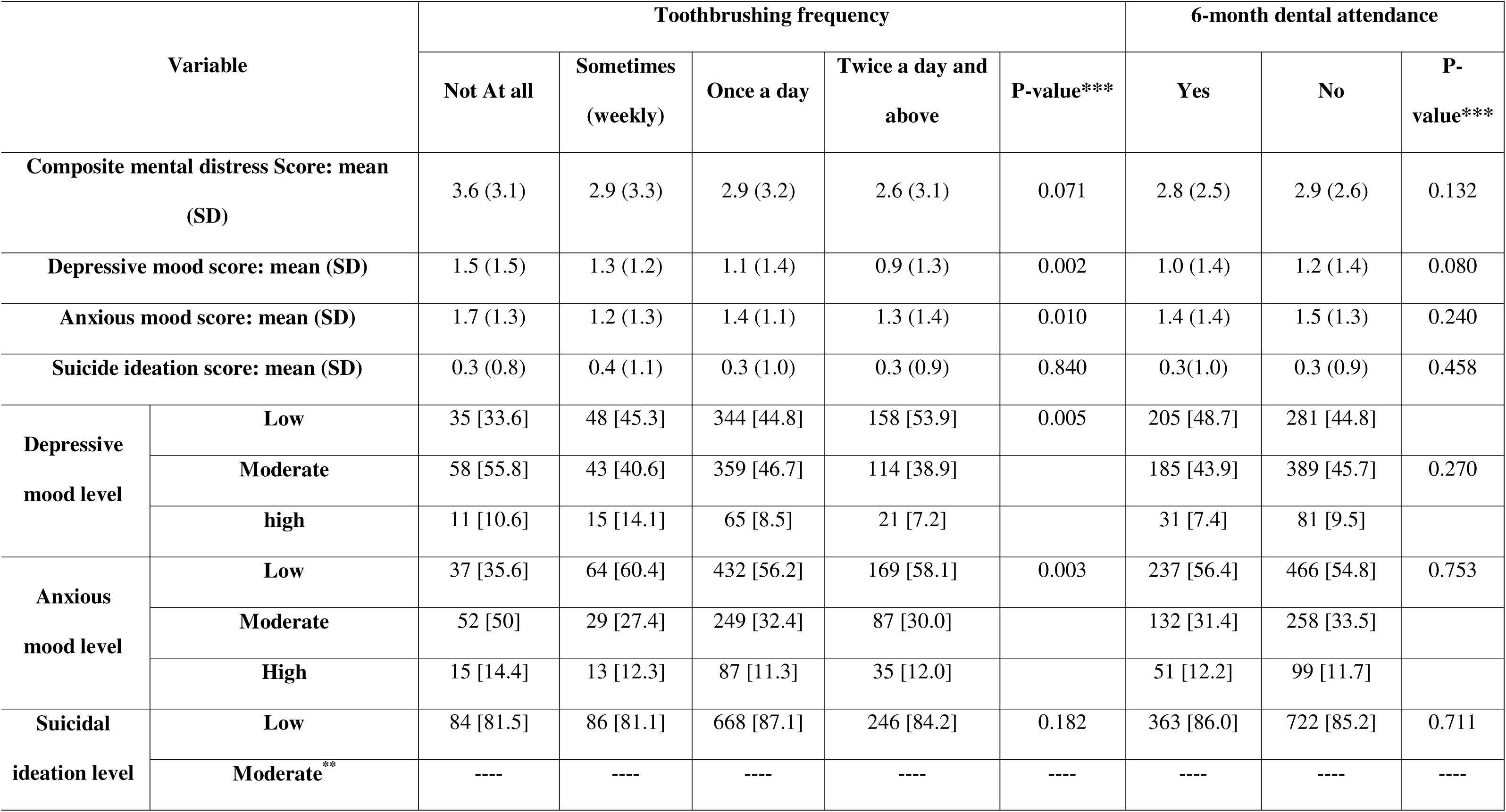
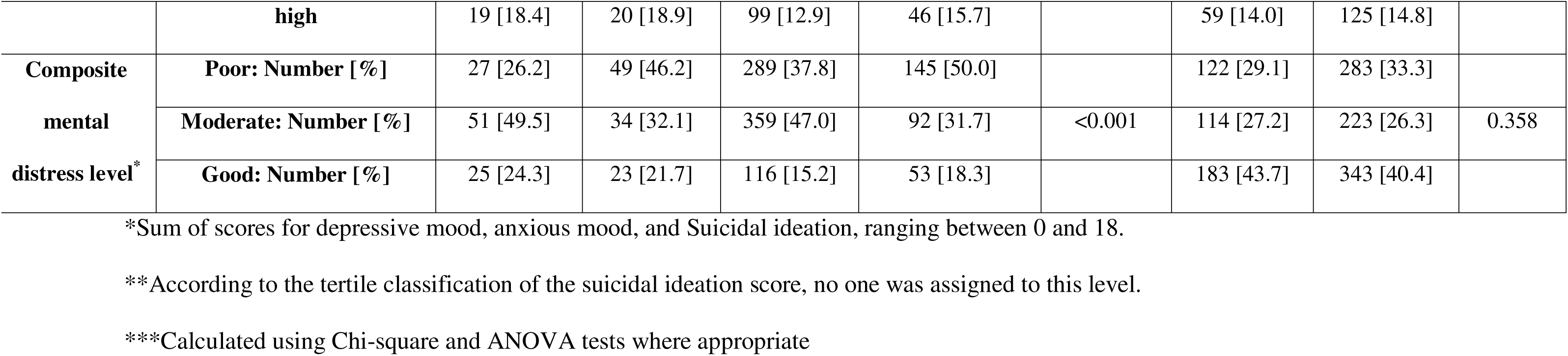
Associations between mental distress indicators, toothbrushing frequency, and 6-month dental attendance among Tehran Province adults (n =1282).

Figure 1 illustrates the relationship between toothbrushing twice a day and above stratified by gender and scores for (A) composite mental distress, (B) depressive mood, (C) anxious mood, and (D) suicidal ideation. A significant, negative, linear association was observed between composite mental distress score and toothbrushing frequency: as scores increased from 0 to 18, the proportion of participants twice a day and above declined, with females consistently toothbrushing twice a day and above more than males (33% vs. 18% at score 0, and 17% vs. 6% at score 18). A similar trend was evident for depressive mood scores, ranging from 0 to 6, with toothbrushing twice a day and above declining from 32% to 22% among females and from 17% to 8% among males. Anxious mood scores showed the same pattern: toothbrushing twice a day and above dropped from 33% to 19% in females and from 17% to 7% in males as scores increased. However, for suicidal ideation scores, the decline in toothbrushing twice a day and above was less pronounced and not statistically significant, with only slight reductions observed for both females (about 30% to about 27%) and males (about 14% to about 11%).

**Figure 1.**
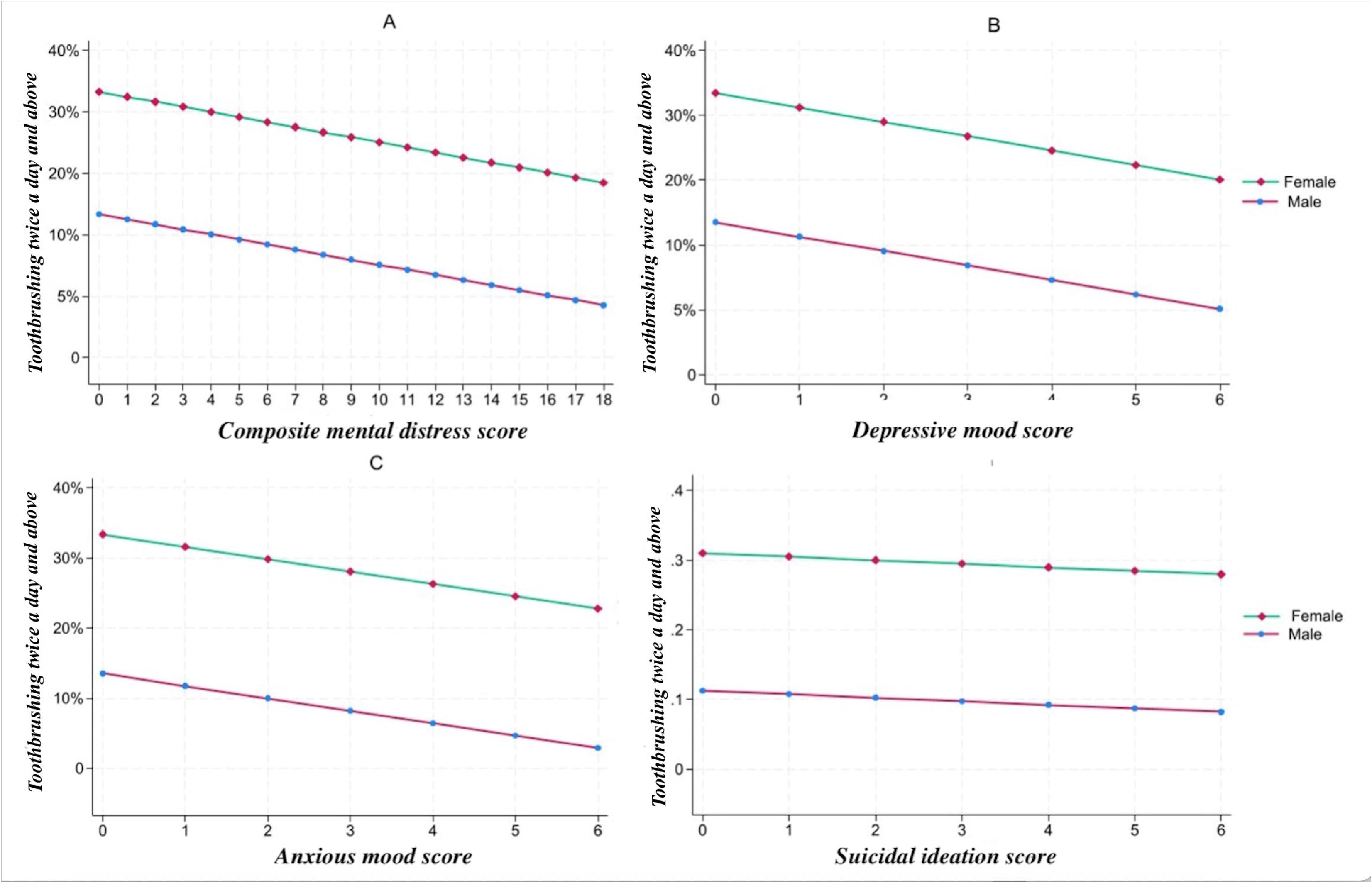
Frequency of toothbrushing twice a day and above across scores for: A) Composite mental distress, B) Depressive mood, C) Anxious mood, and D) Suicidal ideation. Note: all trends were statistically significant (p<0.05) except for the one concerning suicidal ideation.

According to Table 3, after adjusting for confounders, moderate composite mental distress level, versus low, was associated with lower odds of toothbrushing twice a day and above (AOR = 0.52, p < 0.001). Similarly, higher composite mental distress scores were related to a reduced likelihood of toothbrushing twice a day and above (AOR = 0.94, p = 0.019). Moderate and high levels of depressive mood, as well as higher depressive mood scores, were also associated with lower odds of toothbrushing twice a day and above (AOR = 0.65, p = 0.004; AOR = 0.54, p = 0.037; AOR = 0.85, p = 0.006, respectively). Higher anxious mood scores showed a similar pattern (AOR = 0.87, p = 0.021). In contrast, suicidal ideation, whether measured categorically or continuously, was not associated with toothbrushing frequency.

**Table 3.** Mental distress indicators of toothbrushing frequency among Tehran Province adults (n =1282).

| **Mental distress indicator** | **Toothbrushing twice a day and above vs. less than twice a day** |  |  |  |
| --- | --- | --- | --- | --- |
| **Composite mental distress level** | **OR [95%CI]** | **P value** | **AOR<sup>1</sup> [95%CI]** | **P value** |
| Low | Ref |  | Ref |  |
| Moderate | 0.52 [0.38, 0.70] | <0.001 | 0.52 [0.38, 0.72] | <0.001 |
| High | 0.81 [0.56, 0.1.17] | 0.267 | 0.73 [0.45, 1.10] | 0.138 |
| **Composite mental distress score (trend) (0-18)** | 0.95 [0.91, 1.01] | 0.067 | 0.94 [0.89, 0.97] | 0.019 |
| **Depressive mood level** |  |  |  |  |
| Low | Ref |  | Ref |  |
| Moderate | 0.66 [0.50-0.88] | 0.004 | 0.65 [0.48,0.87] | 0.004 |
| High | 0.62 [0.37-1.03] | 0.069 | 0.54 [0.30,0.96] | 0.037 |
| **Depressive mood score (trend) (0-6)** | 0.87 [0.79, 0.96] | 0.010 | 0.85 [0.76, 0.95] | 0.006 |
| **Anxious mood level** |  |  |  |  |
| Low | Ref |  | Ref |  |
| Moderate | 0.83 [0.62-1.11] | 0.217 | 0.78 [0.56,1.07] | 0.127 |
| High | 0.95 [0.63-1.45] | 0.847 | 0.82 [0.52,1.31] | 0.424 |
| **Anxious mood score (trend) (0-6)** | 0.90 [0.82,1.02] | .062 | 0.87 [0.78, 0.97] | 0.021 |
| **Suicidal ideation level** |  |  |  |  |
| Low | Ref |  | Ref |  |
| High | 1.13 [0.79, 1.63] | 0.492 | 1.05 [0.70,1.57] | 0.782 |
| **Suicidal ideation score (trend) (0-6)** | 1.02 [0.90, 1.16] | 0.722 | 0.96 [0.83,1.11] | 0.601 |

1-Adjusted for age, sex, education, residence, smoking, marital status, chronic disease status and health service reception

In unadjusted models, higher depressive and anxious mood scores increased odds of attending dental visits in the past 6 months (OR=1.19, p = 0.005, and OR=1.16, p = 0.016, respectively). However, after adjustment, these associations were no longer significant and none of the other mental distress indicators were significantly related to 6-month dental attendance either (Table 4).

**Table 4.** Mental distress indicators of 6-month dental attendance among Tehran Province adults (n =1282).

| **Mental distress indicator** | **6-month dental attendance (Yes versus No)** |  |  |  |
| --- | --- | --- | --- | --- |
| **Composite mental distress level** | **OR [95%CI]** | **P value** | **AOR<sup>1</sup> [95%CI]** | **P value** |
| Low | Ref |  | Ref |  |
| Moderate | 1.25 [0.86, 1.61] | 0.281 | 1.15 [0.86, 1.65] | 0.280 |
| High | 1.23 [0.93,1.63] | 0.133 | 1.19 [0.84,1.52] | 0.327 |
| **Composite mental score (trend) (0-18)** | 0.97 [0.93, 1.02] | 0.254 | 1.02 [0.93,1.03] | 0.203 |
| **Depressive mood level** |  |  |  |  |
| Low | Ref |  | Ref |  |
| Moderate | 0.88 [0.69, 1.12] | 0.321 | 0.92 [0.7,1.18] | 0.557 |
| High | 0.75 [0.45, 1.11] | 0.136 | 0.73 [0.45,1.19] | 0.212 |
| **Depressive mood score (0-6)** | 1.19 [1.05, 1.34] | 0.005 |  |  |
| **Anxious mood level** |  |  |  |  |
| Low | Ref |  | Ref |  |
| Moderate | 0.91 [0.70, 1.17] | 0.479 | 0.94 [0.72, 1.24] | 0.695 |
| High | 1.01 [0.69, 1.45] | 0.946 | 1.10 [0.74, 1.63] | 0.663 |
| **Anxious mood score (0-6)** | 1.16 [1.03,1.32] | 0.016 | 1.09 [0.96, 1.27] | 0.235 |
| **Suicidal ideation level** |  |  |  |  |
| Low | Ref |  | Ref |  |
| High | 1.03 [0.87,1.23] | 0.711 | 1.08[0.90,1.28] | 0.387 |
| **Suicidal ideation score (0-6)** | 1.0 [0.89,1.13] | 0.814 | 1.06 [0.94,1.21] | 0.309 |

1-Adjusted for age, sex, education, residence, smoking, marital status, chronic disease status and health service reception

## Discussion

The present study reinforced the critical need to view mental health as inseparable from oral health by examining how indicators of mental distress, assessed using rapid mental health screening tools, relate to key oral health behaviors in the Iranian adult population. Findings revealed that higher depressive mood scores, higher anxious mood scores, and moderate and high levels of depressive mood (versus low) were associated with toothbrushing less than twice daily. Similarly, moderate levels of composite mental distress (versus low) and higher mental distress scores were related to toothbrushing less than twice daily. Interestingly, none of the explored mental distress indicators were associated with 6-month dental attendance post-adjustment.

This study has notable strengths, including a large sample, stratified random sampling proportional to size, robust questionnaire development, validated telephone surveys, and comprehensive covariate adjustment. It is also likely the first study to have explored the relationships between rapid mental health screening of depressive and anxious mood, suicidal ideation, and two key oral health behaviors, particularly among Iranian adults, addressing an important gap. Nonetheless, some limitations should be noted. The cross-sectional design precludes causal inference. Self-reported data may be affected by recall or social desirability bias, though logical timeframes and interviewer training were used to mitigate these. Telephone interviews, while cost-effective, may have excluded those without stable access. Finally, the sample, comprising mostly women aged 26–45, individuals with lower SES, and urban residents from southern Tehran province, may have introduced bias into the estimates. Women typically report better oral health behaviors, whereas lower-SES individuals often face challenges with dental attendance, and Tehran is among Iran’s more affluent provinces. Stratified random sampling and statistical adjustments were applied to address this limitation. However, the findings’ generalizability may still be affected.

The relationship observed between moderate and high levels of depressive mood, higher depressive mood scores, and toothbrushing less than twice daily is likely driven by symptoms classically associated with depression, such as fatigue, reduced motivation, anhedonia, and impaired executive function. These symptoms can weaken the individual’s ability or willingness to sustain routine self-care, thereby increasing their likelihood for inadequate toothbrushing [62–64]. This finding broadly aligns with prior studies from Korea, Australia, and a global pandemic-era survey, which demonstrated that depressive symptoms and states, whether chronic, recent, or situational, are associated with suboptimal toothbrushing behavior [25, 61, 65]. However, in contrast, a Turkish study found no associations between the presence and severity of depressive symptoms and the individual’s toothbrushing behavior [66]. This discrepancy might be explained by differences in study populations, as the Turkish study included patients attending a single dental school, who likely differ in demographic characteristics and health behaviors compared to the general adult population in southern Tehran province. Additional variation potentially arises from the measures used to assess depressive symptoms, the operational definitions applied, and categorizations of toothbrushing response options. Therefore, contextual and methodological factors should be considered when investigating depressive symptoms and adult toothbrushing behavior.

Interestingly, higher anxious mood was associated with toothbrushing less than twice daily when analyzed as a continuous variable, but not when categorized. This was likely due to the greater sensitivity of the continuous scale, as converting scores into tertiles to create categorical levels may mask more subtle associations. Anxious mood may compromise toothbrushing through its associations with reduced motivation, cognitive overload, social withdrawal, or executive dysfunction, which can reduce one’s capacity for self-care acts [14, 22, 34]. It can also do so through its relationships with periodontitis, oral ulcers, and bruxism, which can cause pain and thus, make the act of toothbrushing uncomfortable [67–69]. Given the scarce availability of data reporting on general anxiety symptoms and toothbrushing behavior, rather than on social phobia or dental anxiety, this finding had to be compared with findings from a Finnish study, which found that adolescents with moderate or severe generalized anxiety were less likely to toothbrush twice daily [34]. Also, a study in Luxembourg found that adolescents with higher stress were less likely to brush their teeth twice daily [70]. Despite the scarcity of directly comparable literature, contradictory findings still exist. For instance, the previously noted Australian study found no associations between toothbrushing less than twice daily and being diagnosed with anxiety disorders [65]. This discrepancy may stem from methodological differences, such as the Australian study’s use of interviews to diagnose anxiety over the past 12 months within its population birth cohort. Overall, further research is needed to clarify the relationship between anxiety and toothbrushing behavior, as well as the underlying mechanisms, in the Iranian adult population.

The association between moderate composite mental distress (versus low), higher mental distress scores, and toothbrushing less than twice daily was expected, as two components of this variable were independently related to reduced toothbrushing. These associations are likely the result of similar mechanisms proposed for the components, including co-occurrence with symptoms of anhedonia, executive dysfunction, and motivational deficits, which make engaging in self-care difficult [22, 71, 72]. Since “composite mental distress” was a variable uniquely designed for this study, direct comparisons with previous studies were challenging. Nevertheless, comparisons can be broadly drawn with constructs such as ‘psychological distress’ or ‘poor mental health,’ which in studies from the U.S., Caribbean, and Indonesia, captured the combined burden of loneliness, anxiety, suicidal thoughts or attempts, self-rated emotional health, and depression, and were associated with reduced toothbrushing frequency [23, 40, 41]. Further research is needed to clarify the relationship between composite mental distress, as defined in this study and measured using rapid screening tools, and toothbrushing behaviors Interestingly, depressive mood, anxious mood, and composite mental distress were not associated with 6-month dental attendance in this sample, whether measured continuously or categorically. Several factors may explain this lack of association. First, the significant associations between higher depressive and anxious mood scores and lack of dental attendance in the past 6 months became non-significant after adjusting for confounders in the multivariable model. This suggests that the observed associations were largely explained by sociodemographic factors, such as sex, age, education, marital status, and SES, which were themselves significantly related to dental attendance (Table 1). Second, routine health behaviors such as daily toothbrushing may be more sensitive to short-term mental health fluctuations, whereas dental attendance might be more strongly shaped by structural and economic factors. This would be particularly relevant in Iran’s LMIC context, where economic hardships are prevalent and dental visits are typically symptom-driven and largely paid out of pocket [73, 74]. Consistent with these findings, an Australian study found that depressive and anxiety symptoms were unrelated to time since the last dental visit or visit frequency in the past 12 months [65]. In contrast, some U.S. studies reported longer gaps in dental care receipt or higher non-use of oral health services among those with poor mental health [26, 63]. This differences may partly be due to the fact that those studies used different data collection tools, variable definitions, and target populations than the present one. In addition, the U.S. is a high-income country, which contrasts with Iran’s LMIC status and its previously noted challenges. Overall, depressive mood, anxious mood, and composite mental distress might offer limited insight into 6-month dental attendance among adults in southern Tehran unless structural, economic, and contextual determinants are considered.

The null associations between suicidal ideation, toothbrushing frequency, and 6-month dental visits were unexpected. Suicidal ideation often co-occurs with depressive and anxious states [75, 76], which in this study were related to greater odds of toothbrushing less than twice a day. It can also involve anhedonia, apathy, social isolation, hopelessness, and executive dysfunction [36, 77, 78], symptoms known to negatively affect oral health behaviors [14, 22]. Together, these factors suggest a potential link for suicidal ideation that was not observed. Nonetheless, several factors may account for the observed null findings. The low prevalence of suicidal ideation in the sample likely limited statistical power. The 1-month recall period for suicidal ideation may not have aligned with the habitual, long-term nature of toothbrushing, while the previously-noted economic barriers and symptom-driven dental care in Iran could have masked the associations with this behavior [74, 79]. Finally, the cultural stigma surrounding suicidal ideation in Iran may have led to underreporting of suicidal thoughts [80], further attenuating observable effects. All in all, the lack of a statistically significant relationship between suicidal ideation and oral health behaviors in this study does not imply clinical insignificance, as even mild suicidal ideation can meaningfully affect self-care and well-being and thus, warrants attention.

In conclusion, this study revealed that among southern Tehran province adults, higher scores for depressive mood, anxious mood, and composite mental distress, along with moderate and high depressive mood levels and moderate composite mental distress levels were all related to toothbrushing less than twice daily. Suicidal ideation showed no link to toothbrushing frequency, and none of the mental distress indicators were associated with 6-month dental attendance. These findings underscore the need for integrated approaches that bridge mental and oral health services. Oral healthcare providers should be trained to identify depressive mood, anxious mood, or overall mental distress using rapid mental health screening tools, provide targeted behavioral support to promote regular toothbrushing among the individuals identified with these conditions, and refer them to mental health services when appropriate. Likewise, mental healthcare providers should be trained to view toothbrushing behavior as a correlate of depressive and anxious moods and overall mental distress, reinforce it as a therapeutic goal, and refer patients with these conditions to oral health services for behavioral counseling. Effective referral and communication pathways between oral and mental health services are essential for these strategies to succeed. Since suicidal ideation did not serve as a reliable indicator of poor toothbrushing or dental attendance in the present sample, integration efforts between mental and oral healthcare may be more impactful when focused on depressive and anxious moods.

Additionally, to improve 6-month dental attendance, economic triage followed by financial and structural interventions, such as subsidized dental care or expanded insurance coverage, might be more beneficial for this sample. Considering this study’s population-based design while taking into account its limitations, these recommendations can be cautiously applied to other Iranian provinces and similar LMIC contexts.

Future studies should investigate whether composite mental distress is related to other oral health behaviors and identify the mechanisms underlying these associations. They should also further explore the relationship between suicidal ideation and key oral health behaviors. In addition, examining structural and economic barriers to regular dental attendance in LMIC settings and piloting rapid mental health screening tools in dental clinics could provide valuable insights into feasible, integrated care approaches. Longitudinal designs would help determine whether causal relationships exist between rapidly assessed mental distress indicators and oral health behaviors. Finally, recruiting more diverse demographic and geographic groups within Iran and other LMICs, particularly rural populations and broader age ranges, would strengthen the generalizability of findings.

## Data Availability

All data produced in the present study are available upon reasonable request to the authors

## References

1. Glick M, Williams DM, Kleinman DV, Vujicic M, Watt RG, Weyant RJ. A new definition for oral health developed by the FDI World Dental Federation opens the door to a universal definition of oral health. J Am Dent Assoc. 2016;147(12):915–7.

2. Xiang DD, Sun YX, Jiao C, Guo YQ, Fei YX, Ren BQ, et al. Diabetes and periodontitis: the role of a high-glucose microenvironment in periodontal tissue cells and corresponding therapeutic strategies. Stem Cell Res Ther. 2025;16(1):366.

3. Chandra Nayak S, Latha PB, Kandanattu B, Pympallil U, Kumar A, Kumar Banga H. The Oral Microbiome and Systemic Health: Bridging the Gap Between Dentistry and Medicine. Cureus. 2025;17(2):e78918.

4. Barranca-Enríquez A, Romo-González T. Your health is in your mouth: A comprehensive view to promote general wellness. Front Oral Health. 2022;3:971223.

5. Kapila YL. Oral health’s inextricable connection to systemic health: Special populations bring to bear multimodal relationships and factors connecting periodontal disease to systemic diseases and conditions. Periodontol 2000. 2021;87(1):11-6.

6. Hensel ALJ, Nicholson K, Anderson KK, Gomaa NA. Biopsychosocial factors in oral and systemic diseases: a scoping review. Frontiers in Oral Health. 2024;Volume 5 - 2024.

7. Christian B, George A, Veginadu P, Villarosa A, Makino Y, Kim WJ, et al. Strategies to integrate oral health into primary care: a systematic review. BMJ Open. 2023;13(7):e070622.

8. Benzian H GR, Monse B, et al. Promoting Oral Health through Programs in Middle Childhood and Adolescence. In: Bundy DAP SN, Horton S, et al., editor. Child and Adolescent Health and Development. Washington (DC): The World Bank;; 2017.

9. Watt RG, Daly B, Allison P, Macpherson LMD, Venturelli R, Listl S, et al. Ending the neglect of global oral health: time for radical action. The Lancet. 2019;394(10194):261–72.

10. WHO. Iran oral health profile 2022 [updated 2022. Available from: https://cdn.who.int/media/docs/default-source/country-profiles/oral-health/oral-health-irn-2022-country-profile.pdf?utm_source=chatgpt.com.

11. Shoaee S, Masinaei M, Saeedi Moghaddam S, Sofi-Mahmudi A, Hessari H, Shamsoddin E, et al. National and Subnational Trend of Dental Caries of Permanent Teeth in Iran, 1990-2017. Int Dent J. 2024;74(1):129–37.

12. Mohammadpour M, Bastani P, Brennan D, Ghanbarzadegan A, Bahmaei J. Oral health policymaking challenges in Iran: a qualitative approach. BMC Oral Health. 2020;20(1):158.

13. Khoshnevisan MH, Ghasemianpour M, Samadzadeh H, Baez RJ. Oral Health Status and Healthcare System in I.R. Iran. Journal of Contemporary Medical Sciences. 2018;4(3).

14. Z K, Siluvai S, Kanakavelan K, Agnes L, Kp I, G K. Mental and Oral Health: A Dual Frontier in Healthcare Integration and Prevention. Cureus. 2024;16(12):e76264.

15. Johnson A, Kenny A, Ramjan L, Raeburn T, George A. Exploring Oral Health Promotion Among Mental Health Providers: An Integrative Review. International Journal of Mental Health Nursing. 2025;34.

16. Organization WH. Anxiety disorders 2025 [updated 8 September 2025. Available from: https://www.who.int/news-room/fact-sheets/detail/anxiety-disorders.

17. Organization WH. Depressive disorder (depression) 2025 [updated 29 August 2025. Available from: https://www.who.int/news-room/fact-sheets/detail/depression.

18. Harmer B, Lee S, Rizvi A, Saadabadi A. Suicidal Ideation. StatPearls. Treasure Island (FL): StatPearls Publishing Copyright © 2025, StatPearls Publishing LLC.; 2025.

19. Cademartori MG, Gastal MT, Nascimento GG, Demarco FF, Corrêa MB. Is depression associated with oral health outcomes in adults and elders? A systematic review and meta-analysis. Clinical Oral Investigations. 2018;22(8):2685–702.

20. Amarasena N, Luzzi L, Brennan D. Effect of Different Frequencies of Dental Visits on Dental Caries and Periodontal Disease: A Scoping Review. Int J Environ Res Public Health. 2023;20(19).

21. Barbosa A, Pinho RCM, Vasconcelos M, Magalhães BG, Dos Santos M, de França Caldas Júnior A. Association between symptoms of depression and oral health conditions. Spec Care Dentist. 2018;38(2):65–72.

22. Bafageeh F, Loux T. Depression Symptoms Linked to Multiple Oral Health Outcomes in US Adults. JDR Clin Trans Res. 2025;10(1):64–73.

23. Heaton LJ, Santoro M, Tiwari T, Preston R, Schroeder K, Randall CL, et al. Mental Health, Socioeconomic Position, and Oral Health: A Path Analysis. Prev Chronic Dis. 2024;21:E76.

24. Cao R, Lai J, Fu X, Qiu P, Chen J, Liu W. Association between psychological stress, anxiety and oral health status among college students during the Omicron wave: a cross-sectional study. BMC Oral Health. 2023;23(1):470.

25. Folayan MO, Zuniga RA, Ezechi OC, Brown B, Nguyen AL, Aly NM, et al. Associations between Emotional Distress, Sleep Changes, Decreased Tooth Brushing Frequency, Self-Reported Oral Ulcers and SARS-Cov-2 Infection during the First Wave of the COVID-19 Pandemic: A Global Survey. International Journal of Environmental Research and Public Health [Internet]. 2022; 19(18).

26. Okoro CA, Strine TW, Eke PI, Dhingra SS, Balluz LS. The association between depression and anxiety and use of oral health services and tooth loss. Community Dent Oral Epidemiol. 2012;40(2):134–44.

27. Kim YS, Kim HN, Lee JH, Kim SY, Jun EJ, Kim JB. Association of stress, depression, and suicidal ideation with subjective oral health status and oral functions in Korean adults aged 35 years or more. BMC Oral Health. 2017;17(1):101.

28. Fatima Z, Bey A, S.A. Azami, Gupta N, Khan A. Mental depression as a risk factor for periodontal disease: A case–control study. European Journal of General Dentistry. 2016;5:86–9.

29. Mohd Khairuddin AN, Bogale B, Kang J, Gallagher JE. Impact of dental visiting patterns on oral health: A systematic review of longitudinal studies. BDJ Open. 2024;10(1):18.

30. Kumar S, Tadakamadla J, Johnson NW. Effect of Toothbrushing Frequency on Incidence and Increment of Dental Caries: A Systematic Review and Meta-Analysis. J Dent Res. 2016;95(11):1230–6.

31. Joshi S, Suominen AL, Knuuttila M, Bernabé E. Toothbrushing behaviour and periodontal pocketing: An 11-year longitudinal study. Journal of Clinical Periodontology. 2018;45(2):196–203.

32. Minoretti P, Liaño Riera M, Gómez Serrano M, Santiago Sáez A, García Martín Á. Brushing Away the Blues: Self-Reported Oral Hygiene Practices Are Associated With Mild Depressive Symptoms in Airline Pilots. Cureus. 2024;16(5):e60695.

33. Anttila S, Knuuttila M, Ylöstalo P, Joukamaa M. Symptoms of depression and anxiety in relation to dental health behavior and self-perceived dental treatment need. European Journal of Oral Sciences. 2006;114(2):109–14.

34. Pohjola V, Nurkkala M, Virtanen JI. Psychological distress, oral health behaviour and related factors among adolescents: Finnish School Health Promotion Study. BMC Oral Health. 2021;21(1):6.

35. Cebrino J, Portero de la Cruz S. Factors related to depression in adults with oral health problems in Spain (2017 to 2020). Frontiers in Public Health. 2024;Volume 12 - 2024.

36. Folayan MO, Tantawi ME, Oginni O, Oziegbe E, Mapayi B, Arowolo O, et al. Oral health practices and oral hygiene status as indicators of suicidal ideation among adolescents in Southwest Nigeria. PLoS One. 2021;16(2):e0247073.

37. Marques-Vidal P, Milagre V. Are oral health status and care associated with anxiety and depression? A study of Portuguese health science students. J Public Health Dent. 2006;66(1):64–6.

38. Folayan MO, Ibigbami OI, Oloniniyi IO, Oginni O, Aloba O. Associations between psychological wellbeing, depression, general anxiety, perceived social support, tooth brushing frequency and oral ulcers among adults resident in Nigeria during the first wave of the COVID-19 pandemic. BMC Oral Health. 2021;21(1):520.

39. Bahramian H, Mohebbi SZ, Khami MR, Asadi-Lari M, Shamshiri AR, Hessari H. Psychosocial determinants of dental service utilization among adults: Results from a population-based survey (Urban HEART-2) in Tehran, Iran. Eur J Dent. 2015;9(4):542–50.

40. Pengpid S, Peltzer K. Hand and Oral Hygiene Practices among Adolescents in Dominican Republic, Suriname and Trinidad and Tobago: Prevalence, Health, Risk Behavior, Mental Health and Protective Factors. International Journal of Environmental Research and Public Health [Internet]. 2020; 17(21).

41. Santoso CMA, Bramantoro T, Nguyen MC, Nagy A. Lifestyle and psychosocial correlates of oral hygiene practice among Indonesian adolescents. Eur J Oral Sci. 2021;129(1):e12755.

42. Anttila SS, Knuuttila ML, Sakki TK. Relationship of depressive symptoms to edentulousness, dental health, and dental health behavior. Acta Odontol Scand. 2001;59(6):406–12.

43. Bouguezzi A AS, Slim A, Besbes A, Hentati1 H, et al. The Interplay Between Oral Medicine and Mental Health. Journal of Clinical Research and Clinical Trials. 2025;4(1):1–4.

44. Shields RE, Korol S, Carleton RN, McElheran M, Stelnicki AM, Groll D, et al. Brief Mental Health Disorder Screening Questionnaires and Use with Public Safety Personnel: A Review. Int J Environ Res Public Health. 2021;18(7).

45. Vallath AL, Sivasubramanian BP, Ravikumar DB, Lalendran A, Krishnan S, Samanta S, et al. The importance of rapid assessment tools in evaluating mental health in emergency departments among patients with chronic diseases. Front Public Health. 2024;12:1258749.

46. Mulvaney-Day N, Marshall T, Downey Piscopo K, Korsen N, Lynch S, Karnell LH, et al. Screening for Behavioral Health Conditions in Primary Care Settings: A Systematic Review of the Literature. J Gen Intern Med. 2018;33(3):335–46.

47. Neulinger B, Ebert C, Lochbühler K, Bergmann A, Gensichen J, Lukaschek K. Screening tools assessing mental illness in primary care: A systematic review. Eur J Gen Pract. 2024;30(1):2418299.

48. Azizpour Y, Ehsani R, Karimi K, Olyaeemanesh A, Delavari A, Vosoogh-Moghaddam A, et al. Developing a pilot study protocol and lessons from Iran for the integrated and repeated public health surveillance system (IRPHS). Journal of Diabetes & Metabolic Disorders. 2025;24(2):191.

49. Kilic C, Rezaki M, Rezaki B, Kaplan I, Ozgen G, Sağduyu A, et al. General Health Questionnaire (GHQ12 & GHQ28): psychometric properties and factor structure of the scales in a Turkish primary care sample. Soc Psychiatry Psychiatr Epidemiol. 1997;32(6):327–31.

50. NHS. How to keep your teeth clean 2022 [updated 15 February 2022. Available from: https://www.nhs.uk/live-well/healthy-teeth-and-gums/how-to-keep-your-teeth-clean/.

51. Zardak AN, Amini-Rarani M, Abdollahpour I, Eslamipour F, Tahani B. Utilization of dental care among adult populations: a scoping review of applied models. BMC Oral Health. 2023;23(1):596.

52. Marshall-Aiyelawo PhD UMK, Bannick PhD UMR, AuD SB, Gliner M, Ms TM, Muraida DJ, et al. Effect of change in the CG CAHPS survey instrument recall period on patient experience scores on healthcare utilization. Patient Experience Journal. 2019;6:114–23.

53. Le LM, Flores G, Edejer TT-T, Tran TK, Nguyen CTK, Tran DT, et al. Investigating the effect of recall period on estimates of inpatient out-of-pocket expenditure from household surveys in Vietnam. PLOS ONE. 2020;15(11):e0242734.

54. Bhandari A, Wagner T. Self-Reported Utilization of Health Care Services: Improving Measurement and Accuracy. Medical Care Research and Review. 2006;63(2):217–35.

55. Mohamadian R, Khazaie H, Ahmadi SM, Fatmizade M, Ghahremani S, Sadeghi H, et al. The Psychometric Properties of the Persian Versions of the Patient Health Questionnaires 9 and 2 as Screening Tools for Detecting Depression among University Students. Int J Prev Med. 2022;13:116.

56. Wild B, Eckl A, Herzog W, Niehoff D, Lechner S, Maatouk I, et al. Assessing generalized anxiety disorder in elderly people using the GAD-7 and GAD-2 scales: results of a validation study. Am J Geriatr Psychiatry. 2014;22(10):1029–38.

57. Veisy F, Farahani H, Togha M, Gharaee B, Janani L, Aghebati A. Rapid screening for generalized anxiety disorder in patients with migraine. Curr J Neurol. 2021;20(2):102–10.

58. Karimi K, Azizpour Y, Shafaati M, Nejad SM, Ehsani R, Nikfarjam A, et al. Assessing HIV transmission knowledge and rapid test history among the general population in Iran. Scientific Reports. 2025;15(1):4944.

59. Azizpour Y. Developing a pilot study protocol and lessons from Iran for the integrated and repeated public health surveillance system (IRPHS). 2025.

60. Tiwari T, Kelly A, Randall CL, Tranby E, Franstve-Hawley J. Association Between Mental Health and Oral Health Status and Care Utilization. Frontiers in Oral Health. 2022;2.

61. Park SJ, Ko KD, Shin SI, Ha YJ, Kim GY, Kim HA. Association of oral health behaviors and status with depression: results from the Korean National Health and Nutrition Examination Survey, 2010. J Public Health Dent. 2014;74(2):127-38.

62. Stepović M, Stajić D, Rajković Z, Maričić M, Sekulić M. Barriers Affecting the Oral Health of People Diagnosed with Depression: A Systematic Review. Zdr Varst. 2020;59(4):273-80.

63. Tiwari T, Kelly A, Randall CL, Tranby E, Franstve-Hawley J. Association Between Mental Health and Oral Health Status and Care Utilization. Front Oral Health. 2021;2:732882.

64. Karimi P, Zojaji S, Fard AA, Nateghi MN, Mansouri Z, Zojaji R. The impact of oral health on depression: A systematic review. Spec Care Dentist. 2025;45(1):e13079.

65. Kisely S, Najman JM. A study of the association between psychiatric symptoms and oral health outcomes in a population-based birth cohort at 30-year-old follow-up. Journal of Psychosomatic Research. 2022;157:110784.

66. Alkan A, Cakmak O, Yilmaz S, Cebi T, Gurgan C. Relationship Between Psychological Factors and Oral Health Status and Behaviours. Oral Health Prev Dent. 2015;13(4):331–9.

67. Girotti M, Adler SM, Bulin SE, Fucich EA, Paredes D, Morilak DA. Prefrontal cortex executive processes affected by stress in health and disease. Progress in Neuro-Psychopharmacology and Biological Psychiatry. 2018;85:161–79.

68. Park J, Wood J, Bondi C, Del Arco A, Moghaddam B. Anxiety Evokes Hypofrontality and Disrupts Rule-Relevant Encoding by Dorsomedial Prefrontal Cortex Neurons. The Journal of Neuroscience. 2016;36(11):3322.

69. Chemelo VDS, Né YGS, Frazão DR, de Souza-Rodrigues RD, Fagundes NCF, Magno MB, et al. Is There Association Between Stress and Bruxism? A Systematic Review and Meta-Analysis. Front Neurol. 2020;11:590779.

70. Geraets AFJ, Heinz A. The association between adolescent mental health and oral health behavior: The Luxembourg Health Behavior in School-Aged Children study. Frontiers in Dental Medicine. 2022;Volume 3 - 2022.

71. Lee H, Lee E. Subgroups of self-neglect and effects on suicidal ideation among the older adults. Development and Psychopathology. 2024;36(4):1537–45.

72. Dong X, Xu Y, Ding D. Elder Self-neglect and Suicidal Ideation in an U.S. Chinese Aging Population: Findings From the PINE Study. J Gerontol A Biol Sci Med Sci. 2017;72(suppl_1):S76-s81.

73. Zardak AN, Amini-Rarani M, Abdollahpour I, Eslamipour F, Tahani B. Factors associated with dental care utilization among Iranian adult populations based on Anderson model. BMC Public Health. 2025;25(1):280.

74. Rezaei S, Pulok MH, Zahirian Moghadam T, Zandian H. Socioeconomic-Related Inequalities in Dental Care Utilization in Northwestern Iran. Clin Cosmet Investig Dent. 2020;12:181–9.

75. Wiebenga JXM, Dickhoff J, Mérelle SYM, Eikelenboom M, Heering HD, Gilissen R, et al. Prevalence, course, and determinants of suicide ideation and attempts in patients with a depressive and/or anxiety disorder: A review of NESDA findings. Journal of Affective Disorders. 2021;283:267–77.

76. Onaemo VN, Fawehinmi TO, D’Arcy C. Risk of Suicide Ideation in Comorbid Substance Use Disorder and Major Depression. medRxiv. 2022:2022.02.28.22271669.

77. McAuley S, Kangas M. An evaluation of psychological interventions targeting positive affect in the treatment of anhedonia: a systematic review. Clinical Psychologist. 2025;29(2):216–32.

78. Elliott E, Sanger E, Shiers D, Aggarwal VR. Why does patient mental health matter? Part 3: dental self-neglect as a consequence of psychiatric conditions. Dental Update. 2022;49(11):867–71.

79. Jadidfard MP, Zafarmand AH, Shafiei S. Association Between Dental Expenditure and Socioeconomic Status in Iran. Int Dent J. 2024;74(6):1432–7.

80. Hassanian-Moghaddam H, Zamani N. Suicide in Iran: The Facts and the Figures from Nationwide Reports. Iran J Psychiatry. 2017;12(1):73–7.

